# A feasibility study on combining Ayurvedic dietary knowledge and modern nutrition to personalise diets for cancer patients

**DOI:** 10.64898/2026.04.06.26350237

**Authors:** Sonia Velarsan, Shaily Agarwal, N Bhargavi, Prasan Shankar, Megha

## Abstract

**Background:** The European Society for Clinical Nutrition and Metabolism (ESPEN) guidelines on nutrition for cancer patients provides evidence-based dietary recommendations that is routinely deployed by dietitians in oncology settings. Although these can be culturally adapted, they do not adequately address inter-individual variability in treatment-related gastrointestinal symptoms and appetite, issues that increase malnutrition risk in cancer patients. Ayurveda, on the other hand, lacks nutrient-based guidelines but offers a well-grounded dietary framework to assess digestive function and personalise diets. This study investigated the feasibility of combining the two approaches in a clinical setting.

**Methods:** Consenting adult cancer patients diagnosed with any type and stage of cancer were recruited. At baseline, digestive strength, dietary intake, quality and frequency and Patient-Generated Subjective Global Assessment (PG-SGA) score were recorded. Based on this, personalised meal plans (MPs) that combine nutrient guidelines from ESPEN and traditional food concepts to support digestive strength were provided to participants. Follow-ups ranged from 4 weeks – 6 months, at which digestive strength and PG-SGA was noted. To evaluate against a benchmark, meal plans were theoretically constructed using Ayurveda concepts (traditional MP) or ESPEN guidelines (Standard MP) alone.

**Results:** Data is presented for 33 participants, of which 52% had weak digestive strength. Baseline intake averaged 879 ± 329 kcal/day, well below the recommended 1400–1600 kcal/ day level. Traditional MPs improved energy intake but were protein-insufficient, aspects that were addressed in the standard MPs. Diet quantity (1417 ± 145 kcal/day), quality and frequency improved on the integrated MP, with 3 patients achieving optimal digestive strength. Personalised counselling reduced malnutrition risk, as reported by PG-SGA score.

**Conclusion:** Customising dietary advice by overlaying nutrient guidelines with Ayurveda dietary concepts is feasible. The evaluation of digestive strength holds promise for personalising nutrition therapy.

**Trial Registration:** CTRI/2023/07/055657

## INTRODUCTION

Malnutrition, a common feature in cancer patients, is associated with increased morbidity, treatment intolerance and poor prognosis (1,2). The causes for malnutrition are varied and include tumour-induced metabolic alterations, increased systemic inflammation, reduced oral intake and/or impaired nutrient absorption due to the site of cancer and therapy-related side effects (3,4). Dietary counselling, recommendations and monitoring continue to be the primary means of addressing malnutrition in cancer patients. A balanced and nutrient-dense diet can improve treatment outcomes, help minimise side effects and enhance patients’ overall quality of life (1). To prevent and manage malnutrition during cancer treatment, authoritative, practical, evidence-based recommendations have been crafted by the National Cancer Institute and ESPEN. Broadly, these recommend the intake of adequate energy and nutrients, especially protein, to prevent weight loss and muscle wasting (5). While these recommendations are highly useful, they are largely nutrient-centric and do not explicitly address gastrointestinal (GI) issues experienced by a large number of cancer patients (6). GI-related issues include nausea, vomiting, diarrhoea, constipation and mucositis, all of which can influence dietary adherence and effectiveness (7,8). To address both nutrient adequacy and GI issues, diets have to be customised for cancer patients.

Interestingly, Ayurveda, a traditional medical system of India, lays major emphasis on GI phenotypes for diagnosis and prognosis (9,10). In Ayurveda, digestive strength (Sanskrit: agni, roughly translated as the digestive/ metabolic fire) is a cumulative read out of several phenotypes including bowel movements, appetite, and sensations before and after food consumption, such as intestinal discomfort, burping, bloating, and flatulence (11,12). Classical texts of Ayurveda categorise digestive strength under four types: optimal (*sam*ā*gni*), weak (manda□*gni)*, sharp (ti□ks n a□*gni*) or irregular (*vi□am*ā*gni)* [Classical references: Charaka Sa□hitā, Sūtrasthāna, Chapter 27, Verse 341–344, A□□ā□ga H□daya by Vāgbha□a, Sūtrasthāna, Chapter 12, Verse 8–10]. According to the *Charaka Sa□hit*ā (an authoritative Ayurvedic text, *S*ū*trasth*ā*na* 28:4–7), optimal digestive strength supports health and longevity, while disturbances in digestive strength underlie the development of disease. Subsequently, a major goal of traditional medical as well as dietetic therapy is to restore the digestive state to an optimal category (13). In support of this approach, the Ayurvedic dietetic framework provides recommendations on ingredients, food formats and dietary patterns.

Furthermore, ingredients, their combinations and processing methods suggested in Ayurveda to manage digestive strength are a living tradition, thus providing a means to customise diets that are both culturally relevant and feasible. Despite the popularity of Ayurveda-based dietary information and traditional food practices for cancer, there is limited published literature on how traditional dietary concepts and modern nutrition can be combined and evaluated in oncology settings. A key challenge to performing such studies lies in the epistemological gap between modern and traditional dietary concepts, and differences in how ingredients are classified and applied functionally. To overcome this challenge, this study examined the feasibility designing meal plans where ESPEN guidelines were used to meet nutrient adequacy while food choices that are rich in these nutrients, as well as dietary patterns, was refined based on digestive strength (agni) and traditional food concepts (14). The digestive strength and nutrient intake of patients was assessed, based on which meal plans were custom designed for each patient. To understand how the different dietary frameworks (ESPEN vs Traditional) compare, theoretical meal plans using exclusively one approach were designed and analysed, while the integrated meal plan was suggested to patients along with dietary counselling. Although this study was not designed to assess outcomes, Patient-Generated Subjective Global Assessment (PG-SGA), Mid Upper Arm Circumference (MUAC) and weight provided a convenient tool to monitor the effectiveness of counselling patients on the integrated diet.

## PATIENTS AND METHODS

This study was part of a single-centre, observational study being conducted at the Institute of Ayurveda and Integrative Medicine (I-AIM) Healthcare Centre, Bangalore, India. Patient recruitment for dietary studies occurred between July 2023 and January 2025.

### Study Protocol

Ethical approval was obtained from The University of Trans-Disciplinary Health Sciences and Technology Institutional Ethics Committee (Approval Number: TDU/IEC/14/2023/PR53) and placed on record at Clinical Trial Registry of India as CTRI/2023/07/055657.

Patients between 18 and 80 years of age, any gender, diagnosed with any stage of cancer according to the International Classification of Diseases (ICD 11), and presenting at I-AIM were eligible for the study. Participants were excluded if they declined consent or were outside the specified age range. The cohort included both outpatients and inpatients. In addition to dietary counselling, patients were also provided with other traditional therapeutic interventions and in some cases, they continued to be on medications that are part of their conventional treatment.

The interval between pre- and post-counselling assessments varied by patient condition, accessibility and treatment setting. Inpatients were monitored daily after initial counselling to ensure adherence and track progress. Outpatients were monitored at baseline and at scheduled follow-up visits. Weekly phone calls were conducted to reinforce dietary adherence, address concerns, and record any symptoms. Overall, follow-ups ranged from 4 weeks to 6 months. Outpatient follow-ups were suspended if patients refused to cooperate. The last recorded follow-up data points are presented in all figures.

### Digestive strength evaluation

Digestive strength was assessed using a validated Agni assessment questionnaire (15). Although the questionnaire is a self-assessment tool, it was administered by the dietitian to guide patients through the questions and ensure accurate completion. The questionnaire contains a total of eleven questions, and based on the score, digestive strength was categorised as: weak, sharp, irregular or optimal. The categorisation obtained from the questionnaire was independently corroborated by the consulting Ayurvedic physician.

### Dietary evaluation

Four meal plans (MPs) were assessed. Pre-study (Baseline) MP: Pre-counselling information collected via a 24-hour dietary recall at recruitment. Traditional MP: A theoretical meal plan for the patient that would have been advised by the Ayurvedic physician solely using Ayurvedic principles. The dietitian collaborated with the physician to assign portion sizes. Standard MP: A theoretical meal plan for the patient, designed by a certified dietitian using contemporary oncology nutrition practices and based on ESPEN guidelines. Except for adaptation to Indian cuisine, no other traditional food knowledge was considered here. Integrative MP: Post-counselling meal plan suggested to the patient, which was a blend of ayurvedic principles and ESPEN guidelines. (See supplementary tables, Table S2, S3, S4, S5, for example). Nutrient analysis of meal plans was computed using the software “DietCal” version 15.1.1 (Profound Tech Solutions), which is based on values from the Indian Food Composition Tables 2017, National Institute of Nutrition, ICMR (IFCT 2017) (16).

All the meal plans were further assessed using the India-specific Diet Quality Questionnaire (DQQ), which gathers food group (∼29) consumption over the previous 24 hours, via a set of yes/no questions (17). This questionnaire was validated and implemented in 85 countries in the Gallup World Poll in 2021–2023 (18,19). Also, for each meal plan, the frequency of food intake was noted. Food frequency was defined as any instance in which a food or beverage was consumed or advised to be consumed during the day, including items such as tea and coffee.

### Other evaluations

At follow-ups, digestive strength was evaluated using the Agni Questionnaire (15). Nutritional status was assessed using the Patient-Generated Subjective Global Assessment (PG-SGA) (20), a widely used oncology tool to identify malnutrition risk and guide nutritional interventions. Mid-upper arm circumference (MUAC) (21) was measured using a standard non-stretchable tape (Seca 203, Seca GmbH, Germany) (22) at the mid-acromial-radial level, perpendicular to the long axis of the arm, as an indicator of muscle mass and somatic protein reserves. A validated, globally accepted questionnaire to assess cancer-specific Quality of Life, EORTC QLQ-C30 was used.

### Sample Size

This was feasibility study nested in a longitudinal observational study, and no formal sample size calculation was performed.

## RESULTS

### Cohort characteristics

The study cohort is a subset of patients participating in a three-year observational study at the Institute of Ayurveda and Integrative Medicine (I-AIM), Bengaluru. Patients were recruited from July 2023 to January 2025. For this report, data from 33 participants have been included, and their demographic as well as pertinent clinical details are reported in Table 1. The majority of participants were female (57.6%), and most were between 40 and 60 years of age (Table 1). Cancer stage was categorised based on National Comprehensive Cancer Network (NCCN) recommendations, and in this cohort, 42.4% of participants were diagnosed with stage IV disease. In the Indian healthcare setting, conventional oncology services and traditional medicine systems operate largely as parallel and independent pathways. Therefore, the point in the cancer treatment journey at which patients came to I-AIM centre was noted: a majority of patients had completed all required chemotherapy or radiation treatments (42.42%; post-treatment), while 30.3% patients did not wish to seek conventional treatment at all (Ayurveda-only). A smaller percentage (18.8%) were seeking to complement their ongoing conventional treatment with Ayurveda (In treatment), while 9.09% were advised palliative care by their oncologist (Table 1). A majority of patients (63.6%) came to the hospital between 4 months and 2 years after receiving their diagnosis. As this was part of an ongoing observational study, a diverse set of cancer types was observed, with breast cancer being the most common in this cohort (21.2%), which reflects the National Cancer Registry Programme (NCRP) Report 2020 (23). Dietary habits reported by the patients revealed an equal distribution of vegetarian and non-vegetarian participants (42.4%), while 15.2% followed an ovo-lacto vegetarian diet.

**Table 1.**
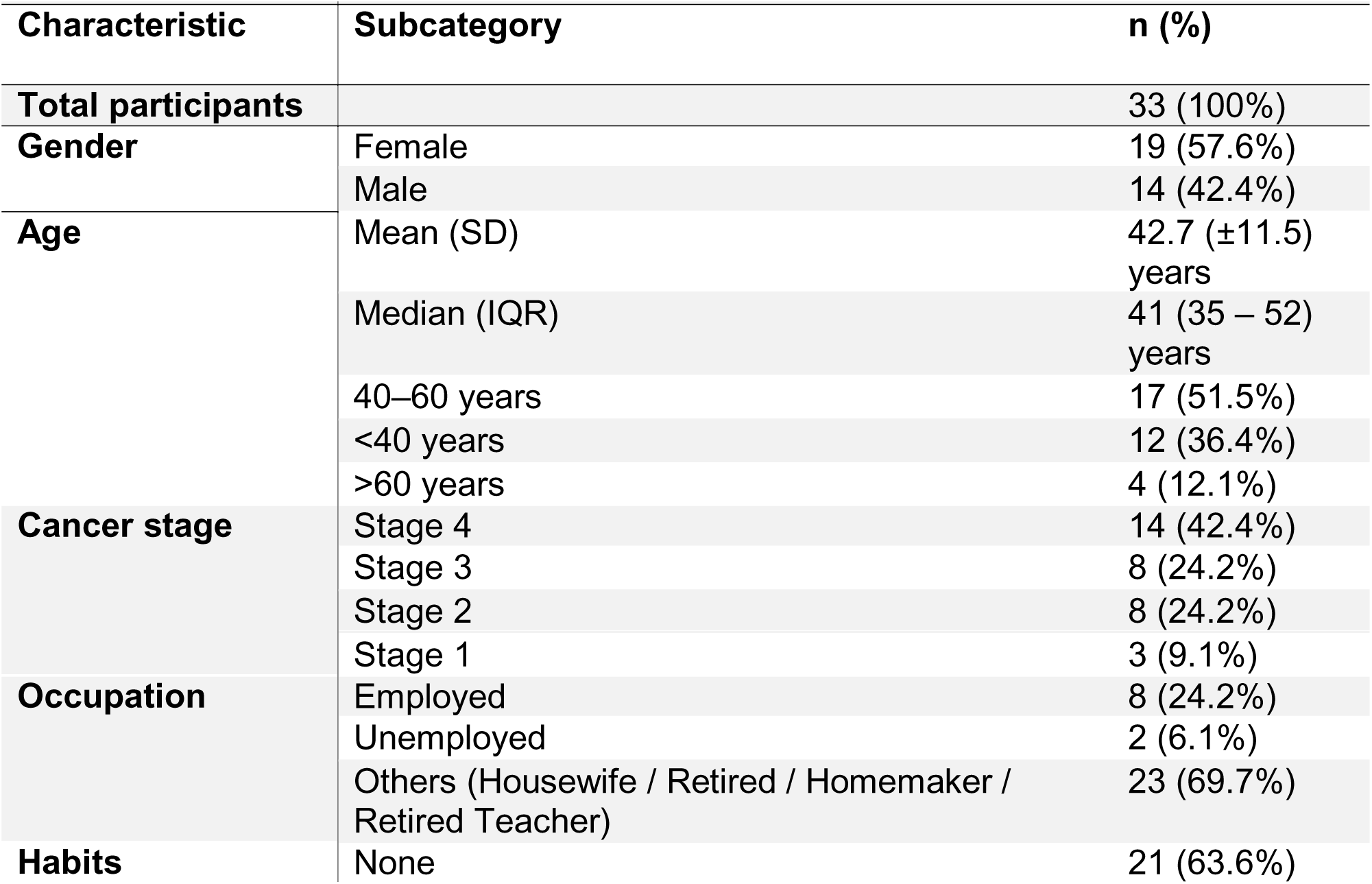
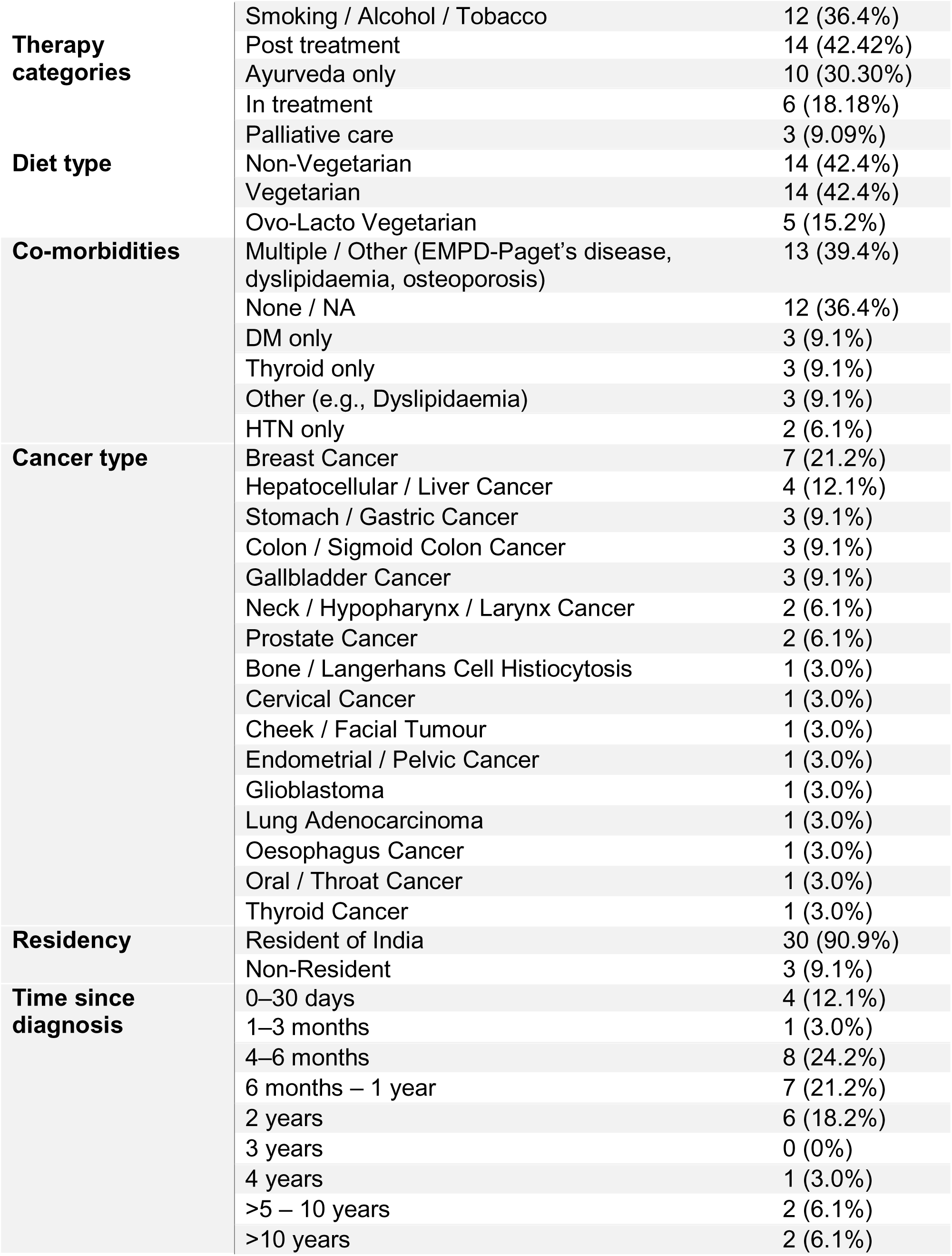
Demographic and Clinical characteristics of study participants (n=33)

### Assessment of Digestive Strength

Within the Ayurvedic framework, assessment of digestive strength (*agni*) forms a central component of dietary planning and clinical evaluation. Using a previously validated questionnaire (15) the digestive strength of patients was categorised. Largely, patients with cancer and under active treatment, or advanced disease pathology, tend to have weak digestive strength. As shown in Figure 1A, at baseline, the majority of patients presented with weak digestive strength (52%, n = 17), followed by irregular digestive strength (39%, n = 13) and sharp digestive strength (9%, n = 3). Notably, no patients exhibited optimal digestive strength at baseline. Post-counselling, the overall distribution pattern among agni types remained largely similar, although 6 patients (18%) demonstrated a change in digestive strength (Supplementary Fig. 1), with 3 of these individuals transitioning to optimal digestive strength. When plotted according to treatment stage, no change in trend for digestive strength was observed (Supplementary Table 1), suggesting that at least in this small heterogeneous cancer cohort, treatment stage and digestive strength are not correlated.

**Figure 1.**
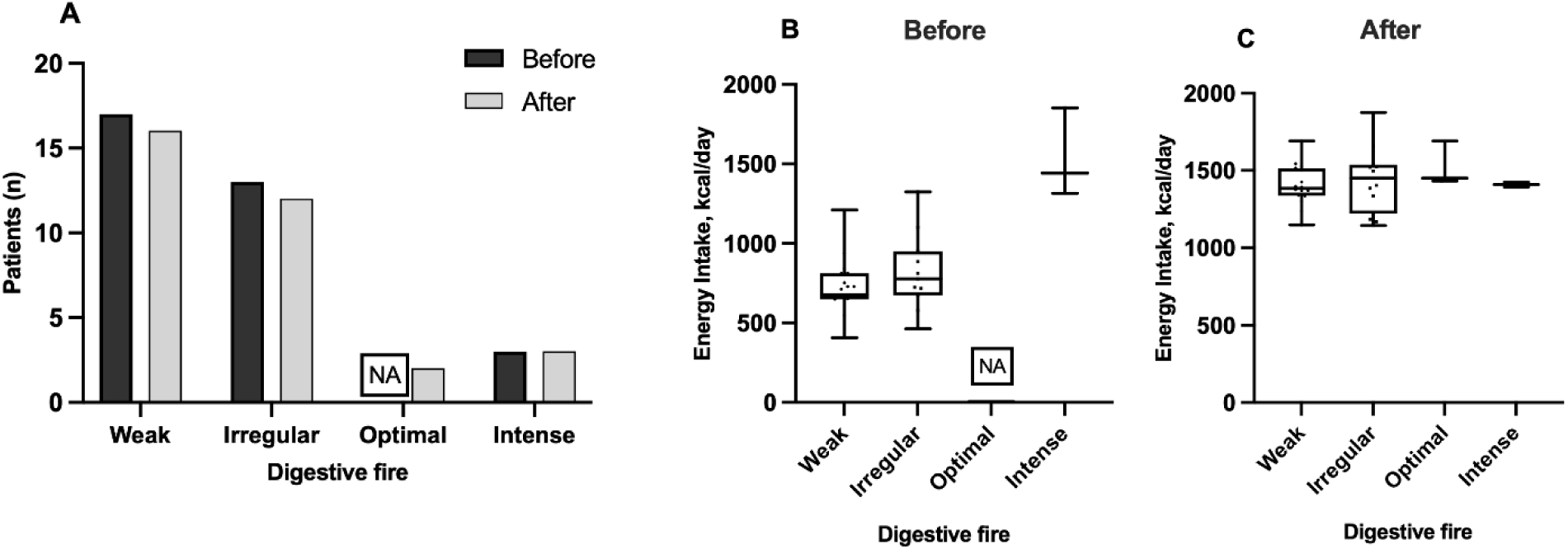
Analysis of digestive strength pre- and post-counselling. (A) Frequency distribution of digestive strength at baseline and follow-up. (B) Digestive strength vs. energy intake at baseline. (C) Digestive strength vs. energy intake at follow-up. ANOVA with Tukey’s post hoc test (α = 0.05) was used. Weak vs. Sharp (*p* < 0.0001). No significant difference was found post counselling (*p* = 0.4718). Significance levels: ns = not significant, *p* < 0.05 (*), p < 0.01 (**), p < 0.0001 (*\*\*\*). NA (not applicable) indicates that no participants fell under the optimal digestive strength pre-counselling.

The desire and capacity for overall food intake may correlate with digestive strength, as feeling hungry on time and desiring to eat are part of the questionnaire. In modern dietetics, the total energy of the consumed foods serves as an indicator of overall food intake. Accordingly, a correlation between 24-hour energy intake and digestive strength was examined (Fig. 1B). At baseline, it was observed that participants with lower energy intake (∼753 kcal/day) primarily exhibited weak digestive strength, while those with higher caloric intake (∼1852 kcal/day) demonstrated intense digestive strength (Fig 1B). However, post-counselling, this association was no longer apparent (Fig. 1C). All patients had similar daily energy consumptions, but continued to exhibit variable digestive strength, suggesting that caloric intake by itself does not capture the traditional concept of digestive strength.

### Nutrient assessment of Integrative meal plans

This study offered the opportunity to evaluate the nutrient and energy differences between meal plans at baseline and those designed using different principles. Data presented here analyses four kinds of 24-hr meal plans (MP) : 1) Pre-study MP: what the patient reported as part of a 24-hour recall at baseline; 2) Traditional MP: what would be recommended by an Ayurvedic physician; 3) Standard MP: dietary recommendations as per ESPEN guidelines; 4) Integrative MP: a diet that combined Ayurvedic principles and ESPEN guidelines. It must be noted that Ayurvedic food advice primarily focuses on ingredients, recipes, and dietary practices; it does not suggest portion sizes based on food groups, as is the norm in contemporary dietetics. This limitation was overcome by the physician and dietician collaborating to derive a reasonable estimate of portion sizes. Moreover, the ayurvedic and standard MPs were a theoretical exercise, hence indicated as grey bars in Figure 2; patients only received the integrative MP during counselling. A sample set of MPs for one patient is provided in the supplemental information.

**Figure 2.**
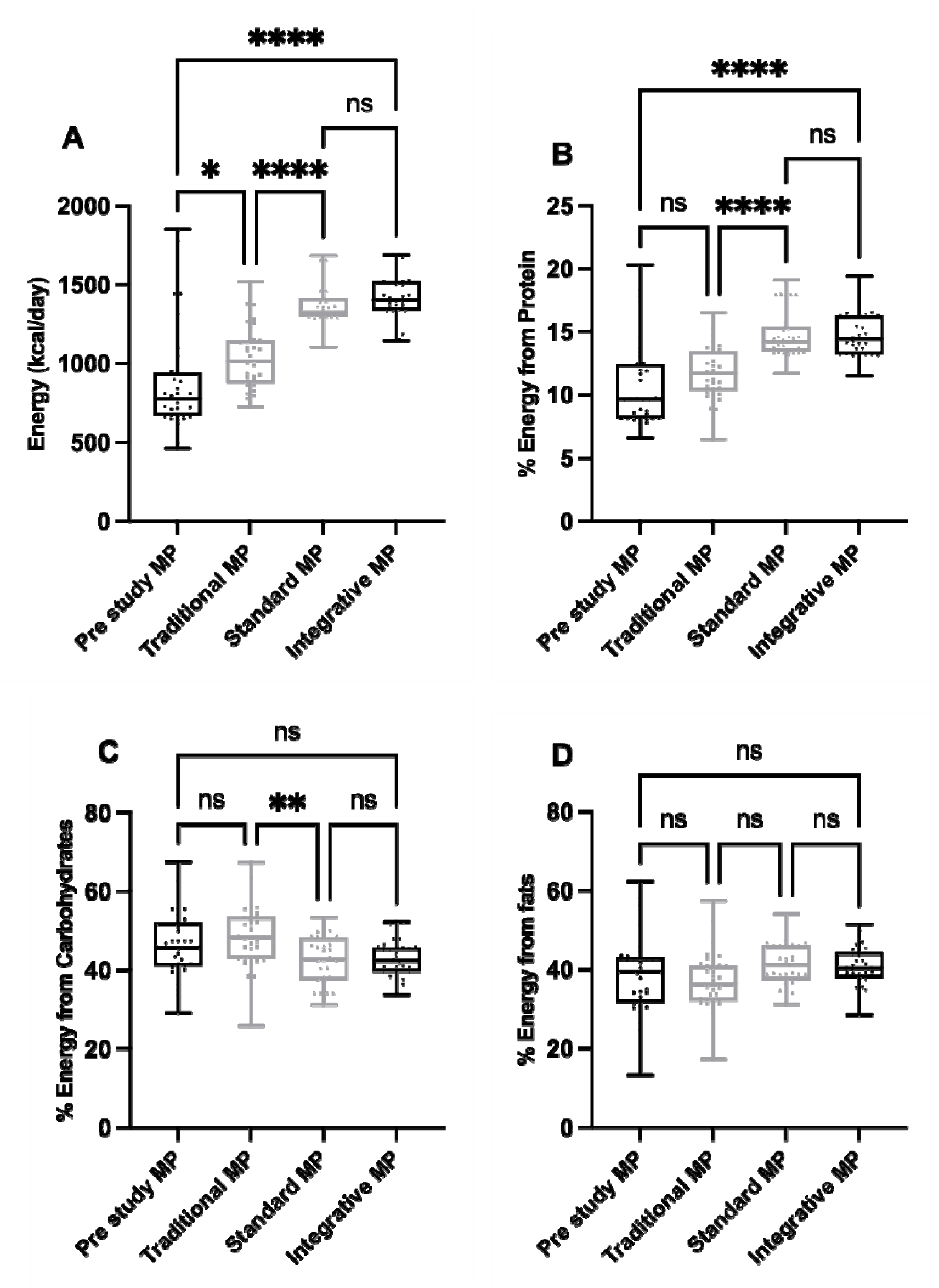
Nutrient evaluation of meal plans (MPs). (A) Total energy intake (kcal/day). Percentage of total calories derived from protein (B), carbohydrate (C) and fat (D). Pre-study MP: what the patient reported as part of a 24-hour recall; Traditional MP: what would be recommended by an Ayurvedic physician; Standard MP: dietary recommendations as per ESPEN guidelines; Integrative MP: a diet that combined Ayurvedic principles and ESPEN guidelines. ANOVA using Tukey’s multiple comparisons test (family-wise alpha = 0.05). Significant differences are denoted as follows: **p < 0.01, ***p < 0.001, ns = not significant.

The average energy intake of the cohort at baseline (Pre-study MPs) was 879 ± 329 kcal/day (Fig. 2A), which is significantly below the ESPEN-recommended intake of 1460-1760 kcal/day (∼ 25–30 kcal/kg/day, which would be expected based on the baseline weight of patients in the study cohort). The theoretical traditional MPs had an increased total energy of 1036 ± 196 kcal/day, but still below recommended levels, while the theoretical standard MPs, formulated following ESPEN guidelines, reached an average energy of 1374.3 ± 127.6 kcal. On the integrative MP, the average total energy increased by 67.8% from the pre-study MP to 1417.6 ± 147.8 kcal/day (p = <0.0001).

With respect to energy derived from macronutrients, a significant difference was observed in the proportion contributed by protein (Fig. 2B) across the four types of meal plans (F = 25.54, p < 0.001). As has been previously reported for cancer patients (24), protein intake in this study too, was found to be low (Pre-study MP = 10.4 ± 3.26%). The ESPEN guidelines recommend a protein intake of 1.0–1.5 g/kg/day (up to 2.0 g/kg/day in severely catabolic or malnourished patients), which corresponds to approximately 13–20% of total energy intake, and up to ∼27% in special cases. Interestingly, protein recommendations on the traditional meal plan were also low (11.7 ± 2.08%). On the standard meal plan, energy from protein increased to 14.4 ± 1.25%. This increase was sustained on the integrative meal plan (14.7 ± 1.80%). Overall, dietary counselling increased protein intake in the diet of cancer patients by approximately 1.5-fold. To increase protein intake, cancer patients are typically advised to consume protein supplements. In this cohort, protein supplement use was reported by 8 participants (24.2%) at baseline, and this increased to 32 participants (97.0%) post-counselling. In the integrative dietary approach, 93% of patients were prescribed protein supplementation to meet their estimated protein requirements. Importantly, the type of supplement prescribed was varied according to the patient’s clinical condition: BCAA-enriched protein formulas were provided for patients with hepatocellular carcinoma, while those with colorectal cancers received semi-elemental formulations. In some patients, protein supplements mixed in diluted buttermilk, instead of milk or water, appeared to improve tolerability and palatability.

Energy from carbohydrates as a percentage of total energy over a 24-hour period was similar across all dietary patterns (Fig. 2C): 46.46 ± 8.01% (pre-study), 48.63 ± 7.55% (traditional), 42.62 ± 6.02% (standard), and 42.97 ± 4.73% (integrative). While a downward trend is observed between pre-study and integrative meal plans, it was not statistically significant. Similarly, energy from fat as a percentage of total energy intake remained statistically unchanged across the four kinds of approaches (Fig. 2D). The pre-study MP had an average fat intake of 38.77 ± 9.76%, while the Traditional MP contained 37.18 ± 7.21%, the standard MP provided 41.29 ± 5.25%, and the integrative MP contained 40.65 ± 5.11%. While an evaluation of the three macronutrient classes offers a convenient strategy for assessing cohort-level changes, they do not provide information on the nutrient sources. As may be observed from the meal plans shared in supplemental tables (Table S2, S3, S4, S5), the source of nutrients varied considerably between diets. Additionally, patients were counselled on appropriate food formats and dietary patterns according to their digestive strength. For those with weak digestive fire, easily digestible and appetizing preparations were advised, such as light nourishing soups incorporating appetite-stimulating spices like dry ginger and long pepper. In such cases, foods lower in energy density but maintaining equivalent total caloric value were recommended to avoid gastrointestinal discomfort while meeting nutritional needs. For example, instead of a whole green gram salad, a green gram soup seasoned with long pepper and rock salt was suggested, as it is lighter on digestion and enhances nutrient assimilation compared to raw or heavy preparations.

To assess qualitative changes in the meal plans, the Diet Quality Questionnaire (DQQ) (19) (Fig. 3A) was used to evaluate dietary diversity. A significant increase in DQQ scores was observed from the pre-study MPs to the traditional to the standard MPs, with the integrative MPs having the highest score of 15 (p < 0.001). Pre-intervention dietary assessment using the Diet Quality Questionnaire (DQQ) showed that participants’ diets were dominated by refined cereals (white rice, wheat roti, idli, dosa), pulses (dal, sambhar), and dairy products (milk, curd). A qualitative assessment of dietary food groups is reported in Table 2. Intake of protective food groups such as green leafy vegetables, vitamin C–rich fruits, nuts, seeds, and lean animal proteins was inadequate. Commonly eaten vegetables included potato, carrot, and pumpkin, while fruit intake was mostly limited to banana and apple. Frequent consumption of sugar-sweetened beverages, fried snacks, biscuits, and confectionery was observed, along with occasional intake of processed foods and soft drinks, contributing to high refined sugar and fat intake. The integrative meal plan included traditional grains, cold-pressed oils, diverse pulses, seasonal vegetables and fruits, and functional foods such as diluted buttermilk, spice teas and light gruels or broths. Specialised medical nutrition formulas, including high-protein, semi-elemental, and disease-specific supplements (e.g., oncology- and hepatic-support formulas) were incorporated based on individual needs. Overall, the post-counselling integrative MP demonstrated a shift from a refined and calorie-dense diet to a nutrient-rich, functionally diverse, and individualised dietary pattern, aligning with both Ayurvedic dietetics and evidence-based nutrition. A low frequency of food intake was observed in participants at baseline, which improved across all three meal plans; following counselling, the average meal frequency increased from 3.69 to 7.27/day on the integrative MP (Fig. 3B).

**Figure 3:**
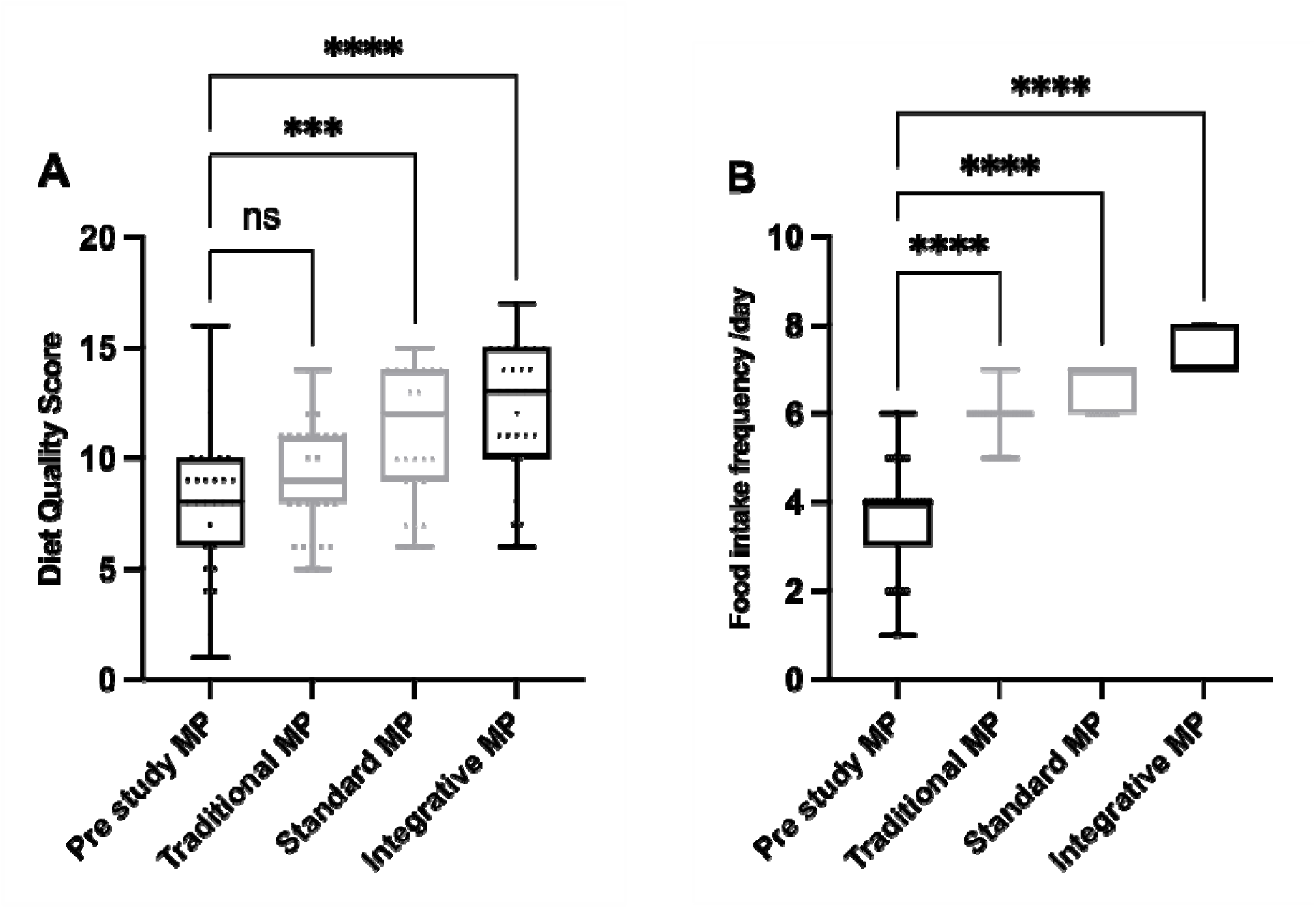
Qualitative changes observed across the meal plans. (A) Diet Quality computed by the DQQ survey. (B) Food intake frequency. ANOVA was performed using Tukey’s multiple comparisons test. Group means: Pre-study = 3.70, Traditional = 5.94, Standard = 6.67, Integrative = 7.27 (n = 33 per group). Significance denoted as ***p < 0.001.

**Table 2.** Comparison of ingredients across food groups between pre-study (24-hour recall) and post-counselling integrative meal plan.

| **Food group** | **Pre-study MP (24-hour recall)** | **Integrative MP (post-counselling)** |
| --- | --- | --- |
| **Cereals** | Bread, pasta, polished rice, wheat roti | Barley, broken wheat, sorghum, finger millet, traditional rice (unpolished rice) |
| **Oils** | Refined oil (sunflower) | Ghee, Cold-pressed sesame and groundnut oil, therapeutic MCT oil |
| **Pulses** | Split pigeon pea, Split Bengal gram, Split yellow gram. | Horse gram, whole green gram |
| **Vegetables** | Beans, carrot, ladies' finger, onion, tomato, pumpkin, snake gourd | Ash gourd, bottle gourd, ridge gourd, snake gourd; seasonal leafy greens |
| **Green leafy vegetables** | Not consumed | Amaranth, fenugreek, moringa, spinach |
| **Milk & milk products** | Buttermilk, cold coffee, cow's milk, curd, flavoured milkshakes, paneer, toned milk | Cow's milk, curd, probiotic drinks, diluted buttermilk, therapeutic whey protein |
| **Fruits** | Apple, banana, orange, guava, papaya, pomegranate (often in juice form) | Amla, pomegranate, banana, guava, orange; reduced packaged fruit juices |
| **Nuts & seeds** | Almonds (occasional), cashews (limited) | Almonds, cashews, flax seeds, walnuts |
| **Animal protein** | Chicken, eggs | Chicken broth, egg whites, fish (mackerel) |
| **Beverages** | Coffee, cold coffee, packaged juices, soups, tea | Amla juice, diluted buttermilk, Spice infused water, gruel, vegetable/lentil broths; reduced tea/coffee |
| **Supplements** | High-protein oral nutrition supplements | Peptide-based enteral formulas, hepatic-specific high-protein, energy-dense formulas, glutamine supplements |
| **Spices & condiments** | Garam masala, green chilli, salt, turmeric | Carom seeds, asafoetida, black pepper, coriander seeds, cumin seeds, curry leaves, dried ginger |
|  |  | powder, fenugreek seeds,<br>mustard seeds and long pepper |

### Select patient outcomes pre- and post-counselling

The study cohort is part of a larger observational study, and large variations existed between cancer types, treatment strategies, and duration of enrolment. Given these limitations, the study focused on outcome measures that could be easily collected and executed. The PG-SGA (Patient-Generated Subjective Global Assessment) tool is a standard measure of the nutritional status of patients with cancer, helping to assess the risk for malnutrition (25). The PG-SGA tool considers multiple factors, including weight changes, dietary intake, symptoms affecting nutrition, and physical function, making it a comprehensive method to track improvements in nutritional status. In the study cohort, a significant change was observed in the PG-SGA score pre and post counselling (Fig. 4A); the average score dropped from 4.5 to 3.3, suggesting a lower risk of malnutrition post counselling on the integrative meal plans and strategies. Interestingly, the highest drop in PG-SGA score was observed in participants in the palliative care category, hinting at the improved quality of life for patients at the end stages (Fig. 4B).

**Figure 4:**
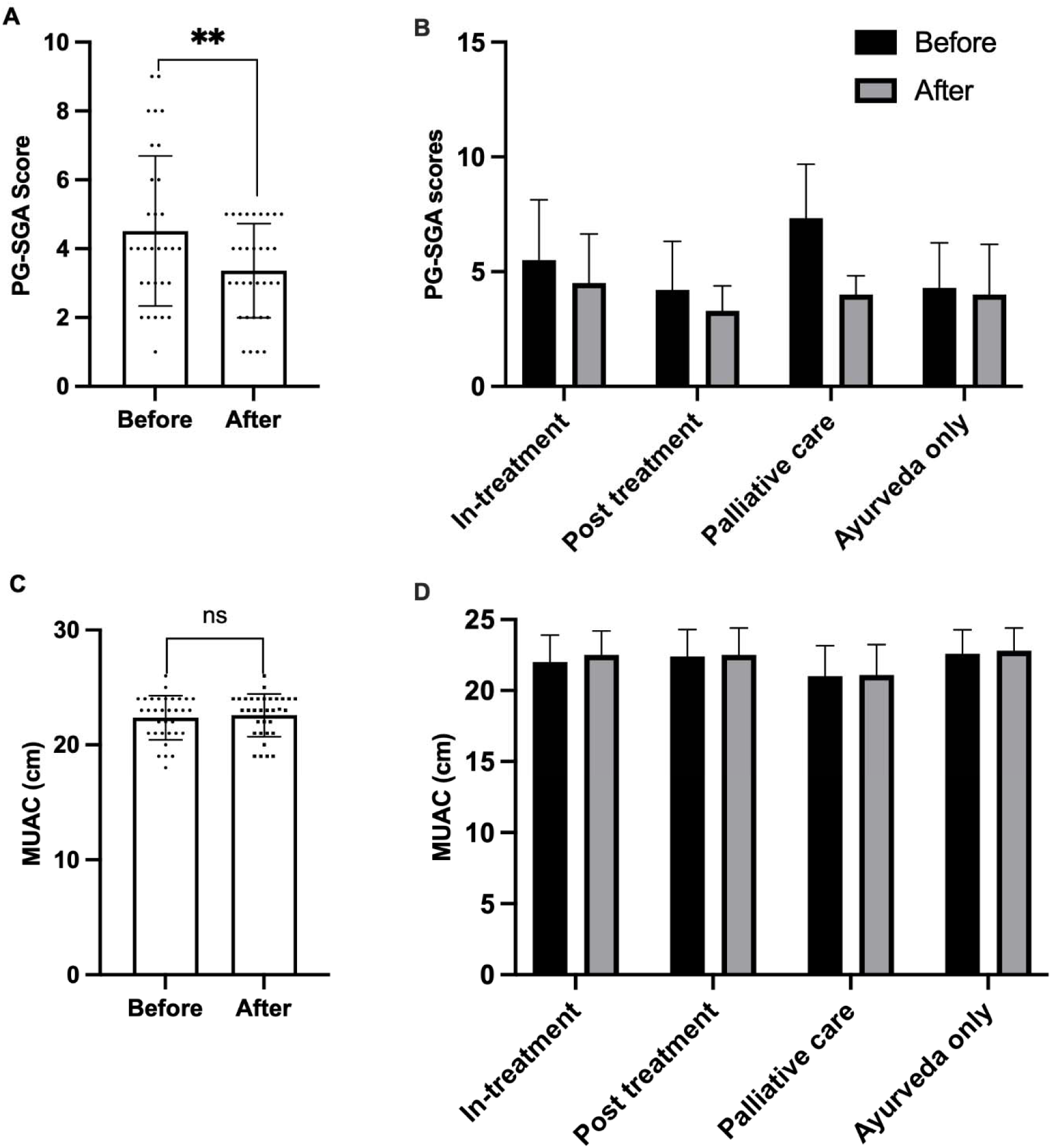
Counselling outcomes of patients. (A) PG-SGA scores at baseline and follow ups (after). Unpaired two-tailed t-test (p = 0.0097). (B) PG-SGA scores stratified by treatment stage at baseline and follow-up (after). (C) MUAC measurements at baseline and follow-up (after). No statistically significant changes were observed in (B) or (C).

Mid-upper arm circumference (MUAC) was measured to assess changes in nutritional status, muscle mass, and subcutaneous fat, as cancer-related malnutrition and cachexia often lead to muscle wasting. It provides a simple, non-invasive indicator of body composition changes, helping to monitor sarcopenia and malnutrition status. (21). The mean MUAC before the intervention was 22.34 cm (SD = 1.94), and after the counselling intervention, it was 22.56 cm (SD = 1.88), reflecting no statistical improvement. Changes in MUAC measurements were stratified based on treatment categories; here, too, no significant differences were observed (Fig. 4D). Weight changes in patients as a percentage increase or decrease over baseline were noted (Supplementary S2). Although not a rigorous marker of nutritional status, weight can be easily tracked by the physician and patient; hence, it was included in the analysis. Weight changes ranged from −5.45% to +5.56% over baseline. On follow-up, 49% (n=17) reported weight gain, 21% (n=7) reported no change, and 30% reported weight loss. Pre- and post-counselling EORTC QLQ-30 Quality of Life (QoL) scores were analysed across three domains: functional scale (S1), symptom scale (S2), and global health scale (S3). Paired t-test analysis demonstrated a significant improvement in the functional scale (S1) (p = 0.03). While changes were observed in the symptom scale (S2) and global health scale (S3), these differences were not statistically significant (Figure S3).

## Discussion

The global cancer burden is projected to exceed 35 million worldwide, a ∼77 % increase from 2022 (26). Conventional treatment relies on surgery, chemotherapy, radiotherapy and/or immunotherapy; diet, as a supportive care, is common to all of them. Despite evidence that diet plays an important role in preventing cancer-related malnutrition and thus improves quality of life, it remains undervalued (27). ESPEN Dietary recommendations, although quite detailed, are generalised. Typically, a dietitian will personalise food choices in the diet plan based on cultural preferences and nutrient targets. This is supported by a recent study amongst Australian oncology dietitians who reported focusing more on increasing energy and protein intake, and less on dietary patterns (28). ESPEN recommendations are not adequate to address gastrointestinal issues, lack of appetite, nausea, lack of taste and other symptoms that reduce dietary intake through diet alone. Ayurveda offers a dietary framework that not only addresses these issues but is also personalised. However, the epistemological gap between modern nutrition and Ayurveda precludes straightforward integration of food choices for diet design. Hence, we set out to assess the feasibility of overlaying traditional food knowledge based on Ayurveda, atop ESPEN guidelines to prepare personalised nutrient-compliant meal plans for cancer patients.

While Ayurveda offers several concepts to consider while designing a diet, here we focus on digestive strength (agni) as a the primary principle. This was chosen because the diagnosis for digestive strength is easy to administer as a questionnaire and in public health nutrition, it would be easier to scale the inclusion of this concept rather than the more complex prakriti-based dietary guidelines. As expected, a majority (52%) of study participants exhibited weak digestive strength, although sharp (9%) and irregular digestive strength (39%) was not uncommon. In Ayurveda, impairment of digestive strength is described as a root cause of systemic dysfunctions and the build-up of waste, concepts that are increasingly paralleled by biomedical descriptions of gut dysbiosis, metabolic inflammation, and their role in carcinogenesis (29–31). However, our data reveals that caution must be exercised in making this assumption.

Based on digestive strength and cultural preferences, patients were presented meal plans with the following main considerations: ingredient choice (e.g., choosing between green gram or horse gram as the protein source), format of consumption (e.g., choosing between a soup vs raw salad) and choice of spices to meet digestive issues such as bloating and constipation. ESPEN guidelines were used to determine the portion sizes in relation to the nutrient composition of the food ingredient. An example of meal plans is presented in Supplementary Table S2-S4. Overall, energy intake and, amongst nutrients, protein intake were less than adequate in cancer patients of this cohort, which is in line with other published literature (32–34). Application of traditional food concepts alone was insufficient to bring energy intake in line with ESPEN guidelines, as they were lower in protein and less diverse. Meal plans based on ESPEN guidelines alone improved protein and energy levels, but had higher levels of energy from carbohydrates. For the study, the strategy of ayurveda-guided food and dietary pattern overlayed on ESPEN nutrient-guidelines resulted in an increase of 67% in energy intake, and 30% increase in protein. The frequency of food consumption increased on all meal plans as compared to baseline. In addition to ingredients, three major changes were observed on the integrated meal plan: an increase in recommendations for soups and drinks with appetising spices such as ginger, fennel; inclusion of two fruits, Indian gooseberry and pomegranate, as juices combined with lentils; and, customised commercial nutrient supplementation to be taken with diluted buttermilk rather than milk (Table 2).

Several limitations of this study are acknowledged. First, the cohort was heterogeneous with respect to cancer type, stage, treatment and follow-up. Hence, digestive strength, PG-SGA and MUAC measurements, are suggestive at best. While weight, PG-SGA and MUAC provide a measure of malnutrition, more sensitive tools, such as body composition analysis (DEXA, BIA), and the use of validated biomarkers of malnutrition (35) will be required to understand if dietary changes translate into changes in nutritional status and lean body mass. Second, patients enrolled in this study were on conventional and traditional medicines, as needed. Dietary counselling and recommended meal plans were part of supportive care. Thus, post-counselling PG-SGA or digestive strength measurements are a sum of medical and dietary inputs. Third, the concept of digestive strength/*agni,* while intuitive, is as yet not fully understood in biological terms. The assessment questionnaire incorporates many elements relating to GI issues that are subjective and rely on patient input alone. The lack of biological markers precludes an independent assessment of the concept as well as corroboration of the changes to digestive state. While no objective biomarker for Agni has yet been established, Singh et al. (2016) report variations in specific aspects pf serum lipid parameters such as LDL/HDL ratio and cholesterol amounts between irregular and optimal digestive strengths (15). Other structured questionnaires based on classical states of *Agni* (*Vishamagni*, *Tikshnagni*, *Mandagni*, *Samagni*) have been proposed (36), which would hopefully encourage more studies to adopt and report on digestive strength as a tool in clinical practice. It must be noted that lack of biological correlates for the concept of “agni” does not preclude it’s utility as a subjective assessment for diet design. However, the lack of large studies demonstrating a positive correlation between agni status and health, or a change in agni status and recovery from illness, may discourage it.

Leveraging traditional medical systems such as Ayurveda, which offer guidelines for personalisation of diets, is an approach that should be investigated as they view ingredients and processing methods from a different functional perspective. For example, rice as an ingredient is considered heavy for digestion and thus, not recommended for an individual with weak digestion. However, if taken as a diluted soup, along with heating spices like pepper and ginger, it is transformed into a food dish that is light and, therefore, suitable for a person with weak digestion. Cellular and molecular studies support the application of herbs such as ashwagandha and turmeric, and herbal mixtures such as triphala in reducing oxidative stress, modulating immunity, and having the potential to improve quality of life in cancer patients (37). This study demonstrates how traditional food knowledge and modern nutrient-based dietary recommendations can be combined. The development of standard methodological approaches to diet design and evaluation is a key first step in order to carry out controlled and rigorous trials to understand if these provide relief from GI symptoms and improve outcomes along the cancer journey. The World Health Organisation has recognised a need for evidence-backed recommendations for the adoption of traditional medicine knowledge (38). It is hoped that this study provides the necessary framework to start investigating dietary interventions that personalise diets based on traditional food knowledge that are also nutrient-compliant.

## Conclusions

Although the frameworks for food classification differ between traditional medical knowledge systems like Ayurveda and modern dietetics, this study demonstrates the feasibility of creating a meal plan that integrates traditional Ayurvedic principles while remaining consistent with modern dietetic recommendations. Because this was a preliminary, small-sample feasibility study, further research with larger cohorts, homogenous cancer-type and stage, and longer follow-up durations is needed to validate the use of digestive strength as a tool to customise diets. It is hoped that the methodological approach presented here would enable clinicians and dieticians to develop better-defined protocols to test the integration of traditional dietary food concepts with modern nutrition for cancer care.

## Data Availability

All data produced in the present study are available upon reasonable request to the authors

## List of Abbreviations

MP: Meal plan
PG-SGA: Patient Generated Subjective Global Assessment
ESPEN: European Society for Clinical Nutrition and Metabolism

## Declarations

### Ethics approval and consent to participate

Ethical approval was obtained from The University of Trans-Disciplinary Health Sciences and Technology Institutional Ethics Committee (Approval Number: TDU/IEC/14/2023/PR53).

### Consent for publication

not applicable

### Availability of data and materials

The datasets used and/or analysed during the current study are available from the corresponding author on reasonable request. Due to patient privacy and ethical considerations, the data are not publicly archived. Researchers interested in accessing the data for academic purposes may contact Dr Megha to request access. Any shared data will comply with applicable legal, ethical, and privacy guidelines.

### Authors’ contributions

**SV**: Investigation, Methodology, Data Curation, Formal Analysis, Writing – original draft, review and editing; **SA**: Investigation, Methodology; **BN**: Investigation; **PS**: Funding acquisition; Project administration; **M**: Conceptualisation, Methodology, Formal Analysis, Supervision, Writing – review & editing; Funding acquisition

## Acknowledgements

We would like to express our sincere gratitude to the study participants. Dr Gurmeet Singh, Dean, TDU, for the support to take up a highly speculative project.

## Author(s’) disclosure (Conflict of Interest) statement(s), even when not applicable

The authors declare that they have no conflicts of interest related to this study.

## Declaration of generative AI and AI-assisted technologies in the manuscript preparation process

During the preparation of this work the authors used ChatGPT in order to improve English language and grammar. After using this tool/service, the authors reviewed and edited the content as needed and take full responsibility for the content of the published article.

## Supplementary Information

### Supplementary figures

**S1.**
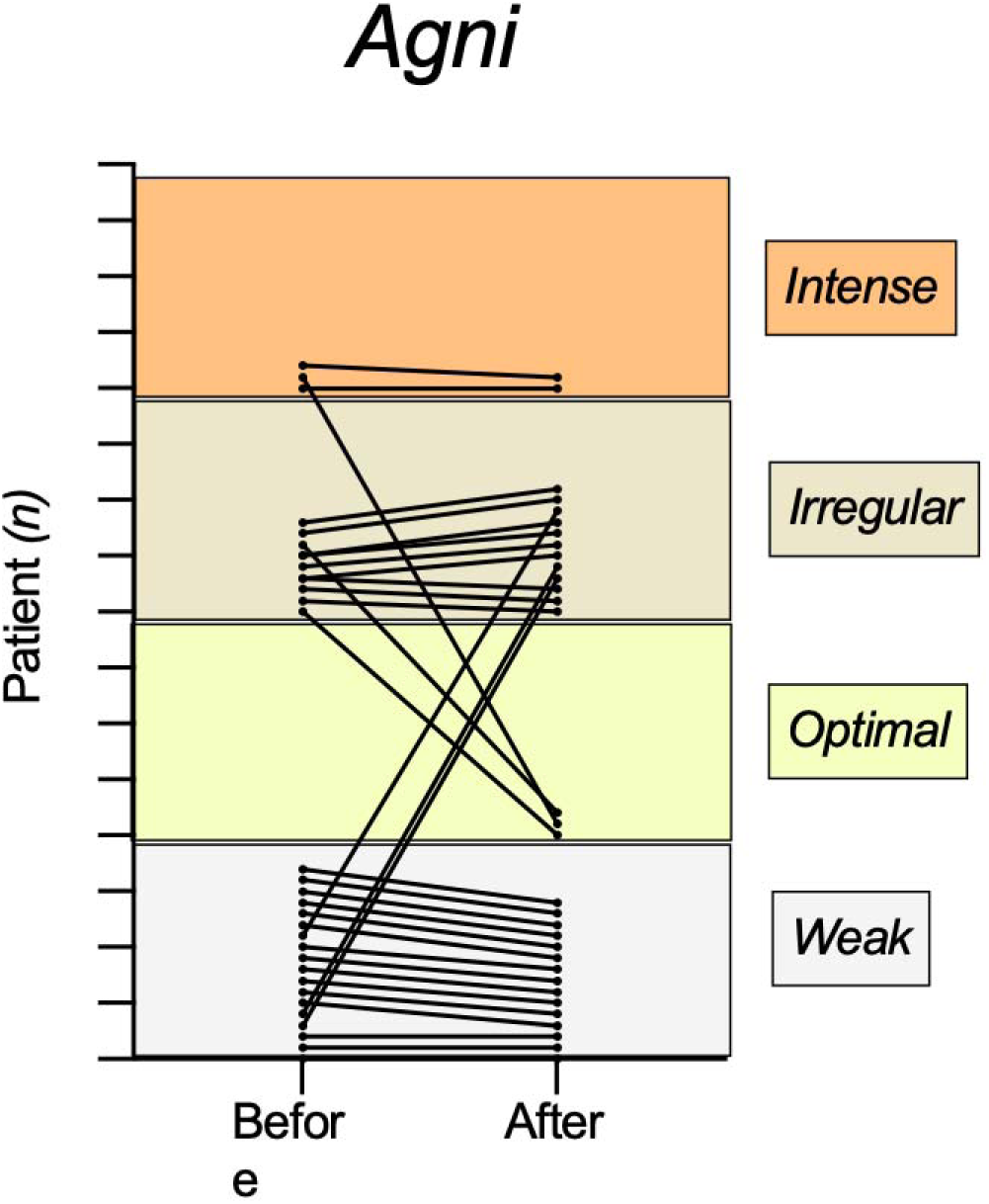
Individual changes in digestive strength between baseline and follow-up. Each line represents the change in Agni category (Weak, Optimal, Irregular, Intense) for a single patient

**S2.**
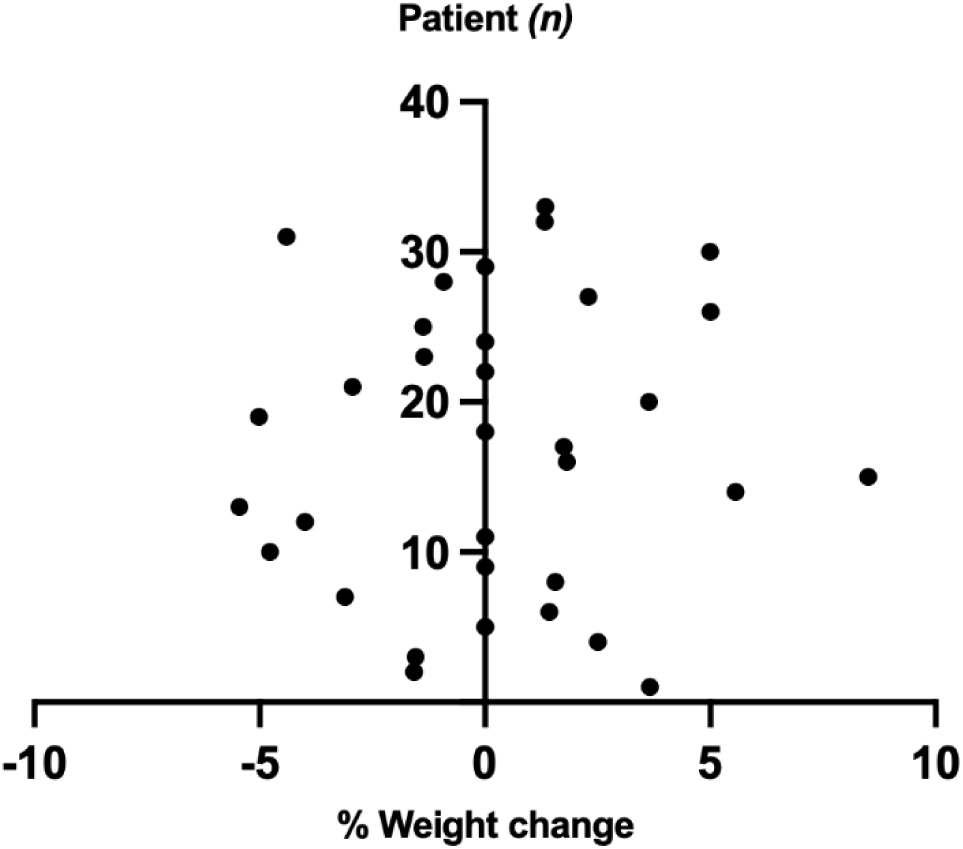
Percentage weight change among study participants. Each dot represents an individual patient. Negative values correspond to a reduction in body weight, 0% denotes no change, and positive values indicate an increase in body weight.

**S3.**
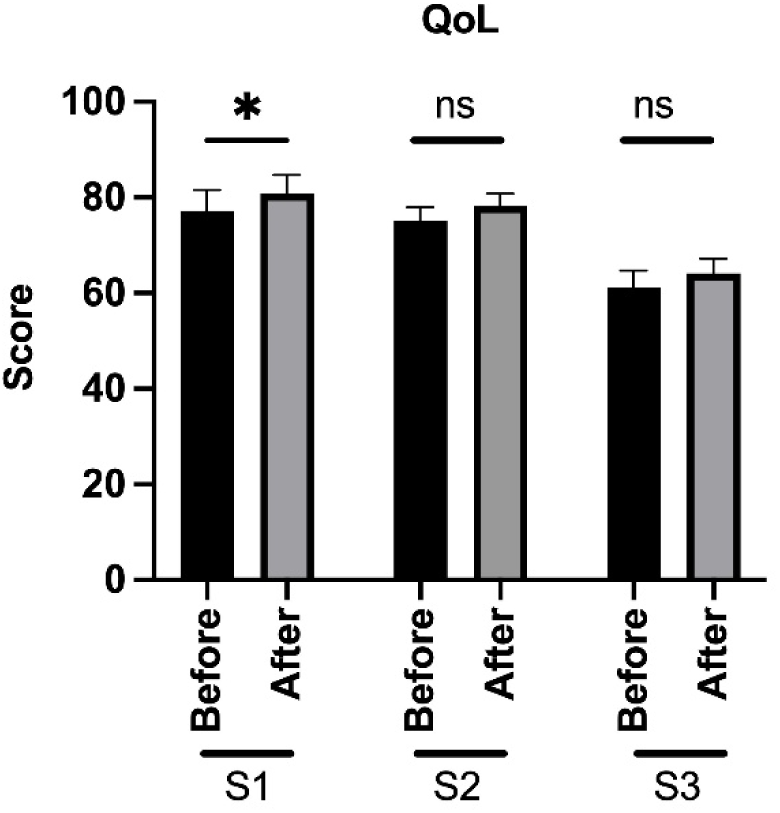
Comparison of Quality of Life (QoL) scores before and after the intervention across three domains: S1 (functional scale), S2 (symptom scale), and S3 (global health scale). A significant improvement was observed in S1 (*p* = 0.03), while changes in S2 and S3 were not statistically significant (ns).

### Supplementary table S1

**Table S1.** Digestive status before and after counselling across different categories.

| Category | Weak (n) |  | Irregular (n) |  | Optimal (n) |  | Intense (n) |  |
| --- | --- | --- | --- | --- | --- | --- | --- | --- |
|  | Before | After | Before | After | Before | After | Before | After |
| **In-treatment** | 3 | 1 | 3 | 4 | 0 | 1 | 0 | 0 |
| **Post-treatment** | 6 | 5 | 4 | 5 | 0 | 1 | 3 | 2 |
| **Palliative care** | 1 | 1 | 2 | 1 | 0 | 1 | 0 | 0 |
| **Ayurveda only** | 8 | 8 | 2 | 2 | 0 | 0 | 0 | 0 |

### Supplementary table S2-S4

Sample set from one patient. MP across the four categories (pre-study, traditional, standard and integrated).

**Table S2:**
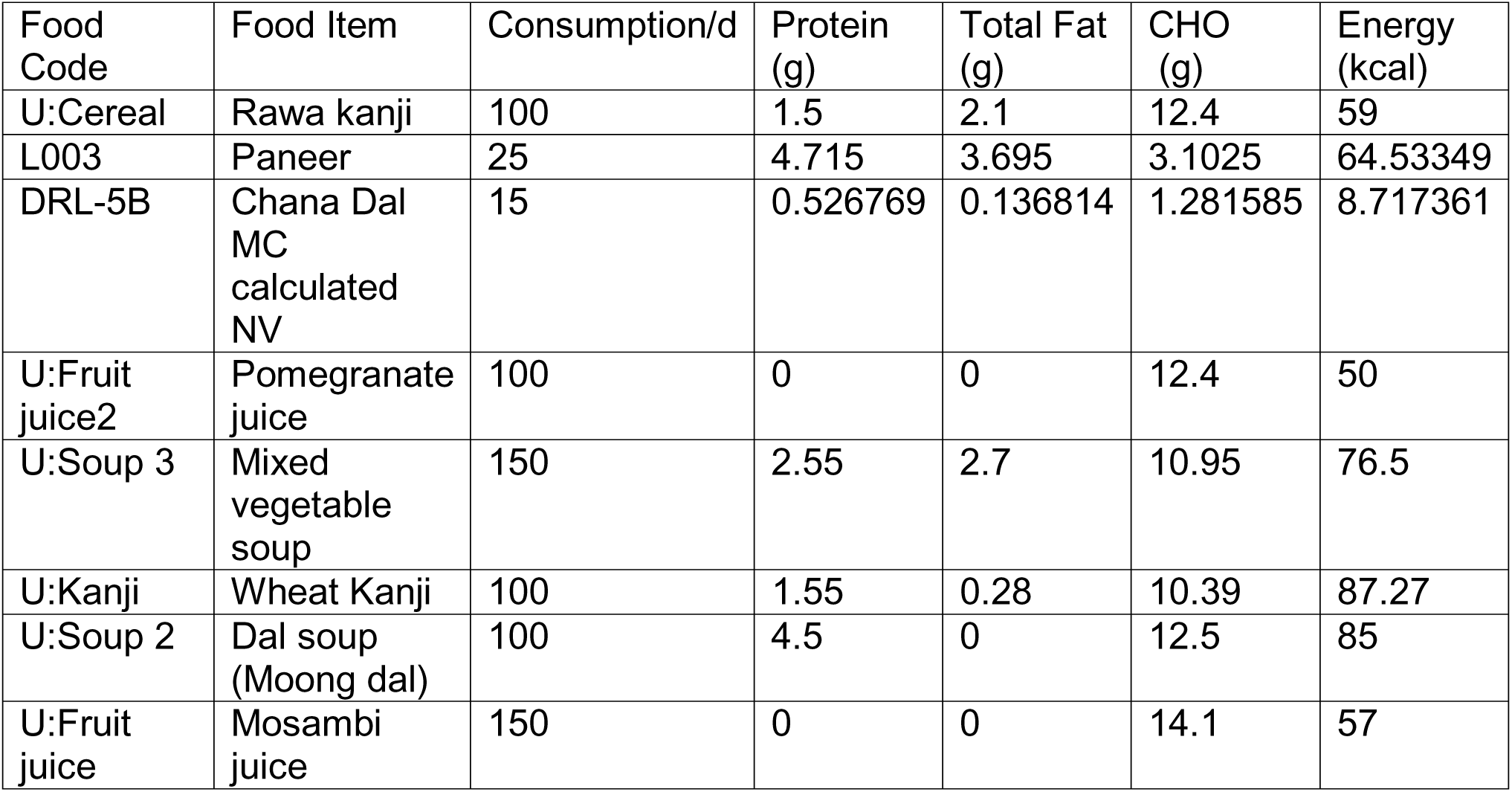
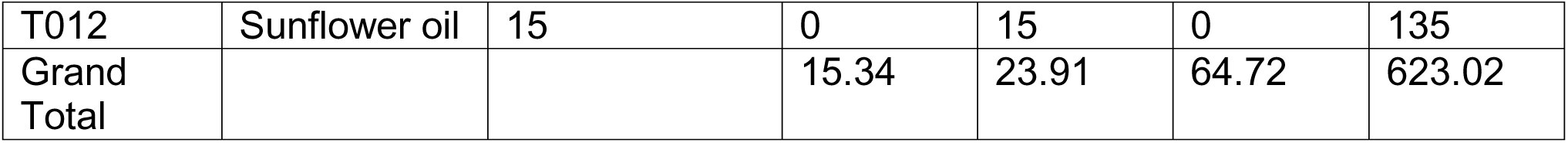
Pre-study MP based on 24hr recall performed on patient.

**Table S3.** Traditional MP based on Ayurvedic principles.

| Food Code | Food Item | Consumption/Day | Protein (g) | Total Fat (g) | CHO (g) | Energy (kcal) |
| --- | --- | --- | --- | --- | --- | --- |
| U:kanji 12 | Rice kanji | 125 | 2.93 | 0.21 | 28.94 | 131.97 |
| U:AIM 6 | Mudgamalaka yusa | 125 | 3.39 | 0.18 | 7.03 | 44.69 |
| U:Cereal | Rava kanji | 100 | 1.50 | 2.10 |  | 59.00 |
| U:AIM 14 | Manda | 150 | 0.30 | 0.02 | 2.93 | 13.34 |
| U:Soup 4 | Vegetable soup (carrot, pumpkin, beans, knolkhol) | 150 | 2.86 | 11.88 | 9.97 | 162.38 |
| U:Fruit juice2 | Pomegranate juice | 145 |  |  | 17.98 | 72.50 |
| U:Soup 2 | Dal soup (Moong dal) | 150 | 6.75 |  | 18.75 | 127.50 |
| U:AIM 9 | Dadimaamalaka (Pomegranate + Amla) | 150 | 0.77 | 0.54 | 8.61 | 38.42 |
| T013 | Ghee | 5 |  | 5.00 |  | 45.00 |
| U:Oils | Cold-pressed gingelly oil | 15 |  | 15.00 |  | 132.60 |
| Grand Total |  |  | 18.49 | 34.93 | 94.20 | 827.40 |

**Table S4.**
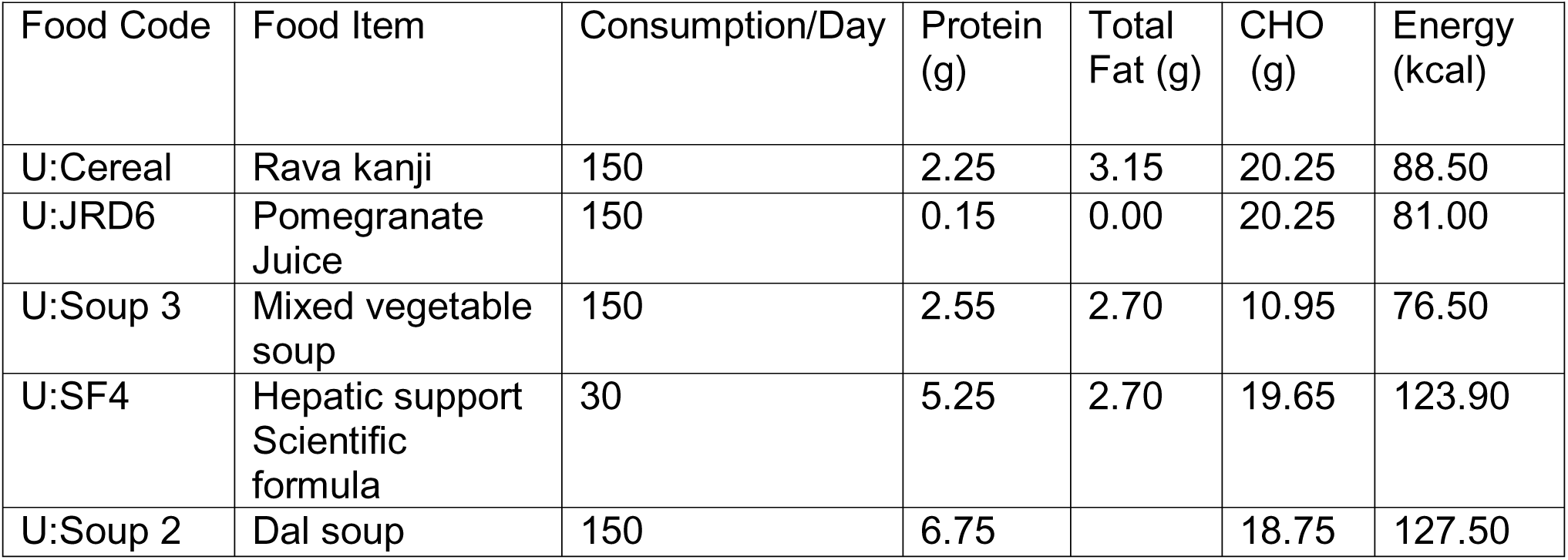
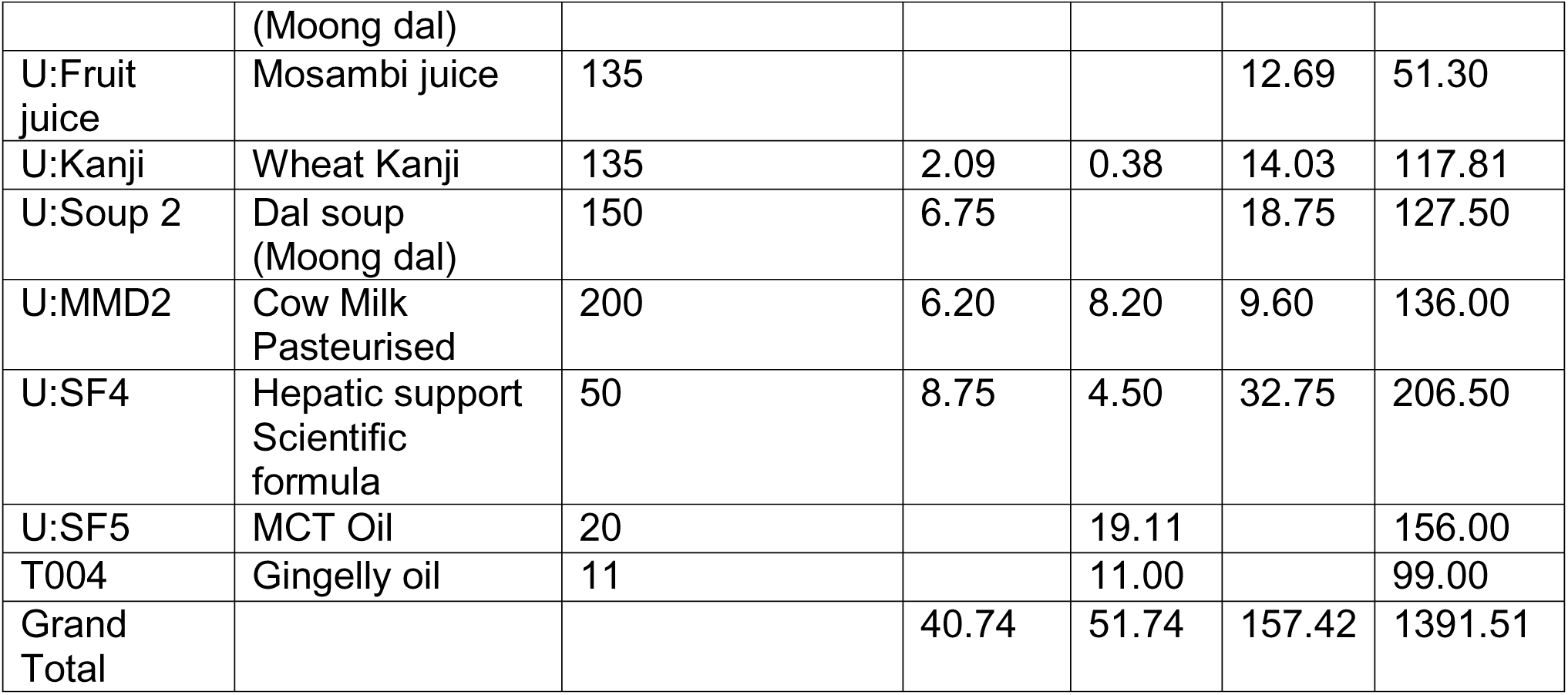
Standard MP designed as per ESPEN guidelines.

| Food Code | Food Item | Consumption/Day | Protein (g) | Total Fat (g) | CHO (g) | Energy (kcal) |
| --- | --- | --- | --- | --- | --- | --- |
| U:Cereal | Rava kanji | 150 | 2.25 | 3.15 | 20.25 | 88.50 |
| U:JRD6 | Pomegranate Juice | 150 | 0.15 | 0.00 | 20.25 | 81.00 |
| U:Soup 3 | Mixed vegetable soup | 150 | 2.55 | 2.70 | 10.95 | 76.50 |
| U:SF4 | Hepatic support Scientific formula | 30 | 5.25 | 2.70 | 19.65 | 123.90 |
| U:Soup 2 | Dal soup | 150 | 6.75 |  | 18.75 | 127.50 |
|  | (Moong dal) |  |  |  |  |  |
| U:Fruit juice | Mosambi juice | 135 |  |  | 12.69 | 51.30 |
| U:Kanji | Wheat Kanji | 135 | 2.09 | 0.38 | 14.03 | 117.81 |
| U:Soup 2 | Dal soup (Moong dal) | 150 | 6.75 |  | 18.75 | 127.50 |
| U:MMD2 | Cow Milk Pasteurised | 200 | 6.20 | 8.20 | 9.60 | 136.00 |
| U:SF4 | Hepatic support Scientific formula | 50 | 8.75 | 4.50 | 32.75 | 206.50 |
| U:SF5 | MCT Oil | 20 |  | 19.11 |  | 156.00 |
| T004 | Gingelly oil | 11 |  | 11.00 |  | 99.00 |
| Grand Total |  |  | 40.74 | 51.74 | 157.42 | 1391.51 |

**Table S5.** Integrative MP: a diet that combined Ayurvedic principles and ESPEN guidelines.

| Food Code | Food Item | Consumption/Day (g) | Protein (g) | Total Fat (g) | CHO (g) | Energy (kcal) |
| --- | --- | --- | --- | --- | --- | --- |
| U:Soup 2 | Dal soup (Moong dal) | 150 | 6.75 |  | 18.75 | 127.50 |
| U:AIM 9 | Dadimamalaka (Pomegranate + Amla) (50+5) | 150 | 0.85 | 0.60 | 9.57 | 42.68 |
| U:SF4 | Hepatic support Scientific formula | 35 | 6.13 | 3.15 | 22.93 | 144.55 |
| U: Kanji | Wheat Kanji | 150 | 2.33 | 0.42 | 15.59 | 130.91 |
| U: AIM18 | Laja tarpana | 150 | 3.45 | 8.55 | 42.32 | 256.95 |
| U: SF4 | Hepatic support Scientific formula | 45 | 7.88 | 4.05 | 29.48 | 185.85 |
| U: Soup 4 | Vegetable soup (carrot, pumpkin, beans, knolkhol) | 150 | 2.86 | 11.88 | 9.97 | 162.38 |
| U: SF5 | MCT Oil | 20 |  | 19.11 |  | 156.00 |
| U:Soup 2 | Dal soup (Moong dal) | 160 | 7.20 |  | 20.00 | 136.00 |
| U: AIM 3 | Mudganna (3 rice: 1 split green-gram) | 100 | 3.37 | 0.22 | 13.12 | 69.58 |
| DRC-9 | Idli Homemade<br>(Rice to Dal 5:1) | 50 | 2.05 | 0.15 | 13.91 | 66.83 |
| DRL-15A | Bood Kumbalakai<br>Sambar (Ash<br>Gourd Sambar) | 100 | 1.87 | 3.66 | 6.10 | 66.16 |
| Grand<br>Total |  |  | 44.72 | 51.80 | 201.73 | 1545.39 |

## References

1. Arends J, Bachmann P, Baracos V, Barthelemy N, Bertz H, Bozzetti F, et al. ESPEN guidelines on nutrition in cancer patients. Clin Nutr Edinb Scotl. 2017 Feb;36(1):11–48. doi:10.1016/j.clnu.2016.07.015 PubMed PMID: 27637832.

2. Caccialanza R, De Lorenzo F, Gianotti L, Zagonel V, Gavazzi C, Farina G, et al. Nutritional support for cancer patients: still a neglected right? Support Care Cancer Off J Multinatl Assoc Support Care Cancer. 2017 Oct;25(10):3001–4. doi:10.1007/s00520-017-3826-1 PubMed PMID: 28710645.

3. Fearon K, Strasser F, Anker SD, Bosaeus I, Bruera E, Fainsinger RL, et al. Definition and classification of cancer cachexia: an international consensus. Lancet Oncol. 2011 May;12(5):489–95. doi:10.1016/S1470-2045(10)70218-7 PubMed PMID: 21296615.

4. Laviano A, Meguid MM, Inui A, Muscaritoli M, Rossi-Fanelli F. Therapy insight: Cancer anorexia-cachexia syndrome--when all you can eat is yourself. Nat Clin Pract Oncol. 2005 Mar;2(3):158–65. doi:10.1038/ncponc0112 PubMed PMID: 16264909.

5. Bozzetti F, Mariani L. Defining and classifying cancer cachexia: a proposal by the SCRINIO Working Group. JPEN J Parenter Enteral Nutr. 2009;33(4):361–7. doi:10.1177/0148607108325076 PubMed PMID: 19109514.

6. Alzoubi Z, Loman BR. Nutrition Interventions in the Treatment of Gastrointestinal Symptoms during Cancer Therapy: A Systematic Review and Meta-analysis. Adv Nutr. 2025 Jul 22;16(9):100485. doi:10.1016/j.advnut.2025.100485 PubMed PMID: 40706958; PubMed Central PMCID: PMC12489516.

7. Kipouros M, Vamvakari K, Kalafati IP, Evangelou I, Kasti AN, Kosti RI, et al. The Level of Adherence to the ESPEN Guidelines for Energy and Protein Intake Prospectively Influences Weight Loss and Nutritional Status in Patients with Cancer. Nutrients. 2023 Sep 30;15(19):4232. doi:10.3390/nu15194232 PubMed PMID: 37836516; PubMed Central PMCID: PMC10574131.

8. Xie SW, Huang JX, Qu HM, Feng ZG, Wang XY, Du ZG, et al. Nutritional management adherence via an ePRO platform in patients with cancer: a machine learning model study. eClinicalMedicine. 2025 Jul 1;85. doi:10.1016/j.eclinm.2025.103330 PubMed PMID: 40620610.

9. Ayurveda and Traditional Chinese Medicine: A Comparative Overview - Patwardhan - 2005 - Evidence-Based Complementary and Alternative Medicine - Wiley Online Library [Internet]. [cited 2025 Oct 28]. Available from: https://onlinelibrary.wiley.com/doi/10.1093/ecam/neh140

10 Integrating ayurvedic medicine into cancer research programs part 1: Ayurveda background and applications - PubMed [Internet]. [cited 2025 Oct 29]. Available from: https://pubmed.ncbi.nlm.nih.gov/36543691/

11. (PDF) Bridging ancient wisdom and modern science: Exploring the symbiotic relationship between Agni and manas in Ayurveda and contemporary perspectives [Internet]. [cited 2025 Oct 28]. Available from: https://www.researchgate.net/publication/380207525_Bridging_ancient_wisdom_and_modern_science_Exploring_the_symbiotic_relationship_between_Agni_and_manas_in_Ayurveda_and_contemporary_perspectives

12. Lad V. Textbook of Ayurveda. Ayurvedic Press; 2002. 376 p.

13. Sakarge DM, Pandey DV, Rahangdale DD, Turkar DA. Conceptual review on Agnimandya with reference to Ajeerna (indigestion) and its Ayurveda management. J Ayurveda Integr Med Sci. 2020 Feb 29;5(01):191–3. doi:10.21760/jaims.v5i01.836

14. Manohar R, Kessler CS. Āyurveda’s Contributions to Vegetarian Nutrition in Medicine. Forsch Komplementarmedizin 2006. 2016;23(2):89–94. doi:10.1159/000445400 PubMed PMID: 27159979.

15. Singh A, Singh G, Patwardhan K, Gehlot S. Development, Validation, and Verification of a Self-Assessment Tool to Estimate Agnibala (Digestive Strength). J Evid-Based Complement Altern Med. 2017 Jan;22(1):134–40. doi:10.1177/2156587216656117 PubMed PMID: 27381899; PubMed Central PMCID: PMC5871217.

16. DietCal [Internet]. [cited 2025 Oct 13]. Available from: http://dietcal.in/

17. (PDF) Measuring what the world eats: Insights from a new approach [Internet]. [cited 2025 Oct 13]. Available from: https://www.researchgate.net/publication/364461545_Measuring_what_the_world_eats_Insights_from_a_new_approach

18. Herforth AW, Wiesmann D, Martínez-Steele E, Andrade G, Monteiro CA. Introducing a Suite of Low-Burden Diet Quality Indicators That Reflect Healthy Diet Patterns at Population Level. Curr Dev Nutr. 2020 Dec;4(12):nzaa168. doi:10.1093/cdn/nzaa168 PubMed PMID: 33344879; PubMed Central PMCID: PMC7723758.

19. Uyar BTM, Talsma EF, Herforth AW, Trijsburg LE, Vogliano C, Pastori G, et al. The DQQ is a Valid Tool to Collect Population-Level Food Group Consumption Data: A Study Among Women in Ethiopia, Vietnam, and Solomon Islands. J Nutr. 2023 Jan;153(1):340–51. doi:10.1016/j.tjnut.2022.12.014 PubMed PMID: 36913471.

20. PG-SGA©. Pt-Global [Internet]. 2014 Mar 27 [cited 2025 Oct 13]. Available from: https://pt-global.org/pt-global/

21. Zhuang CL, Zhang FM, Xu HX, Weng M, Yao Y, Zhou FX, et al. Reference values of low body mass index, mid-upper arm circumference, and calf circumference in cancer patients: A nationwide multicenter observational study. Nutr Burbank Los Angel Cty Calif. 2022;99-100:111688. doi:10.1016/j.nut.2022.111688 PubMed PMID: 35594630.

22. seca [Internet]. [cited 2025 Oct 13]. seca 203 - Ergonomic circumference measuring tape with extra Waist-To-Hip-Ratio calculator (WHR). Available from: https://www.seca.com/en_in/products/all-products/product-details/seca203.html

23. Report of National Cancer Registry Programme 2020 [Internet]. [cited 2025 Oct 24]. Available from: https://www.ncdirindia.org/All_Reports/Report_2020/default.aspx

24. Dingemans AM, van Walree N, Schramel F, Soud MYE, Baltruškevičienė E, Lybaert W, et al. High Protein Oral Nutritional Supplements Enable the Majority of Cancer Patients to Meet Protein Intake Recommendations during Systemic Anti-Cancer Treatment: A Randomised Controlled Parallel-Group Study. Nutrients. 2023 Dec 7;15(24):5030. doi:10.3390/nu15245030 PubMed PMID: 38140289; PubMed Central PMCID: PMC10745925.

25. Azevedo MD, de Pinho NB, de Carvalho Padilha P, de Oliveira LC, Peres WAF. Clinical usefulness of the patient-generated subjective global assessment short form© for nutritional screening in patients with head and neck cancer: a multicentric study. ecancermedicalscience. 2024 Feb 1;18:1662. doi:10.3332/ecancer.2024.1662 PubMed PMID: 38439803; PubMed Central PMCID: PMC10911671.

26. Global Cancer Facts & Figures [Internet]. [cited 2025 Oct 21]. Available from: https://www.cancer.org/research/cancer-facts-statistics/global-cancer-facts-and-figures.html

27. Muscaritoli M, Arends J, Bachmann P, Baracos V, Barthelemy N, Bertz H, et al. ESPEN practical guideline: Clinical Nutrition in cancer. Clin Nutr. 2021 May;40(5):2898–913. doi:10.1016/j.clnu.2021.02.005 PubMed PMID: 33946039.

28. Curtis AR, Kiss N, Livingstone KM, Daly RM, Ugalde A. Exploring dietitians’ practice and perspectives on the role of dietary patterns during cancer treatment: A qualitative study. PloS One. 2024;19(5):e0302107. doi:10.1371/journal.pone.0302107 PubMed PMID: 38743744; PubMed Central PMCID: PMC11093385.

29. Patwardhan B, Mutalik G, Tillu G. Integrative Approaches for Health: Biomedical Research, Ayurveda and Yoga. Academic Press; 2015. 383 p.

30. Sethi JK, Hotamisligil GS. Metabolic Messengers: tumour necrosis factor. Nat Metab. 2021 Oct;3(10):1302–12. doi:10.1038/s42255-021-00470-z PubMed PMID: 34650277.

31. Kumari S, Srilatha M, Nagaraju GP. Effect of Gut Dysbiosis on Onset of GI Cancers. Cancers. 2025 Jan;17(1):90. doi:10.3390/cancers17010090

32. Steindorf K, Clauss D, Rötzer I, Tjaden C, Hackert T, Wiskemann J. Nutrition Intake and Nutrition Status of Pancreatic Cancer Patients: Cross-Sectional and Longitudinal Analysis of a Randomized Controlled Exercise Intervention Study. Nutr Cancer. 2022;74(10):3492–500. doi:10.1080/01635581.2022.2077382 PubMed PMID: 35608567.

33. Gabrielli CP, Steemburgo T. Adequate calorie and protein administration via enteral nutrition may contribute to improved 30-day survival in patients with solid tumors at nutritional risk. Clin Nutr ESPEN. 2024 Feb;59:279–86. doi:10.1016/j.clnesp.2023.12.014 PubMed PMID: 38220387.

34. Subih HS, Al-Shwaiyat EA, Al-Bayyari N, Obeidat BS, Abu-Farsakh F, Bawadi H. Dietary Intake Is Not Associated with Body Composition nor with Biochemical Tests but with Psychological Status of Cancer Patients Receiving Chemotherapy. Nutrients. 2023 Dec 13;15(24):5087. doi:10.3390/nu15245087 PubMed PMID: 38140346; PubMed Central PMCID: PMC10746082.

35. Wunderle C, Urbach K, Buchmueller L, Randegger S, Kaegi-Braun N, Laviano A, et al. Biomarkers for individualized nutritional therapy in disease-related malnutrition: a narrative review. Am J Clin Nutr. 2025 Sep;122(3):671–9. doi:10.1016/j.ajcnut.2025.07.009 PubMed PMID: 40675491.

36. Eswaran HT, Kavita MB, Tripaty TB, Shivakumar. Formation and validation of questionnaire to assess Jā harāgni. Anc Sci Life. 2015;34(4):203–9. doi:10.4103/0257-7941.159829 PubMed PMID: 26283805; PubMed Central PMCID: PMC4535068.

37. Jha SK, Singh N, Shanker OR, Antil I, Baghel JS, Huddar V, et al. A review on integrative approaches in oncology: bridging ayurvedic medicine and modern cancer therapeutics. Front Nat Prod. 2025 Aug 22;4. doi:10.3389/fntpr.2025.1635197

38. Global traditional medicine strategy 2025-2034 [Internet]. [cited 2026 Jan 2]. Available from: https://www.who.int/publications/i/item/9789240113176

